# The treatment of scaphoid non-unions with partial and total scaphoid in titanium custom-made 3D-printed prostheses and reconstruction of scapholunate interosseous ligament: a preliminary study

**DOI:** 10.1101/2021.09.05.21262363

**Authors:** Alessio Cioffi, Mario Igor Rossello

**Author notes:** **Corresponding Author** Alessio Cioffi, MD, Department of Orthopaedic Surgery (DICHIRONS), University of Palermo, Via del Vespro, 90100, Palermo, Italy.

## Abstract

The treatment of sequelae of scaphoid fractures is still one of the unsolved problems of hand surgery. Between January 2019 and July 2020 in the Hand Surgery Department of San Paolo’s Hospital in Savona (Italy) 9 partial scaphoid in titanium custom-made 3D-printed prosthesis have performed in 9 patients (all men) with an average age of 27.5 years (minimum 18-maximum 37 years). The aim of the study is to evaluate over time the clinical-functional and radiographic results of the wrists treated with partial and total scaphoid titanium custom-made 3D-printed prostheses and reconstruction of SLIL.

## INTRODUCTION

The treatment of sequelae of scaphoid fractures is still one of the unsolved problems of hand surgery. These types of lesions, if diagnosed late, result in non-unions, with or without necrosis. The natural history is a radioscaphoid osteoarthritis, the alteration of the biomechanic complex of the radiocarpal joint, which ranges from the instability of intercalated segment (VISI or DISI) to carpal collapse (so called SNAC wrist)^1^. Many treatments are available to reconstruct the scaphoid or to prevent and treat the related sequelae: like bone grafts (free or pedunculated when the pseudoarthrosis is isolated, without carpal collapse phenomena), partial or total resection of the scaphoid in association with mediocarpal arthrodesis, corpectomy of the first row, total wrist replacement or total wrist arthrodesis.^2-3-4^ When is not possible to save the scaphoid and there are no mediocarpal or radiocarpal arthrosis signs or collapse, a total scaphoid prosthetic replacement can be considered for the treatment. While partial replacement is indicated when non-union has led to isolated aseptic necrosis of the proximal pole of the scaphoid.^5^

The history of scaphoid prostheses is long and studded with successes and failures. It started with the first scaphoid prosthesis made in vitallium in 1945, followed by the acrylic implant in 1950. ^6-7^ Then Swanson’s prosthesis of 1962 was constructed in Silistic, but it developed many problems of “silicon synovitis” which in some cases produced massive degeneration of the wrist, passing through the same in titanium which demonstrated good performances even at long term controls^8^, but never reached a wide diffusion. Later Bioprofile^®^ introduced a pyrocarbon adaptive proximal scaphoid implant (APSI), based on a completely different concept of adaptation of the carpus to the implant, and not in an anatomical reconstruction^9^. The simplicity of the prosthesis allowed a good diffusion of the technique, with good results reported by many authors in the short and medium term, but with frequent phenomena of collapse of the carpus and penetration of the implant into the long-term neighboring bone segments.^10^ Since 2018, thanks to the development of powder technology and the custom made implants engineering, it is possible to build total or partial scaphoid prostheses created “made to measure” thanks to 3D CT reconstructions of the single wrist. The indications for this procedure are very compelling: scaphoid destruction, unsuitable for a reconstruction with grafting techniques, good wrist stability and absence of SNAC-wrist condition, as demonstrated by carpal height and radio-scaphoid angle measurements, absence of degenerative changes in the radial scaphoid facet and/or other carpal bones^11^. These new prostheses are built to allow the reconstruction of the scapholunate interosseous ligament (SLIL), an important element of stability of the implant and biomechanics of the carpus ^5^. The aim of the study is to evaluate over time the clinical-functional and radiographic results of the wrists treated with partial and total scaphoid titanium custom-made 3D-printed prostheses and reconstruction of SLIL.

## MATERIALS AND METHODS

### Partial scaphoid prosthesis

Between January 2019 and July 2020 in the Hand Surgery Department of San Paolo’s Hospital in Savona (Italy) 9 partial scaphoid in titanium custom-made 3D-printed prosthesis have performed in 9 patients (all men) with an average age of 27.5 years (minimum 18-maximum 37 years). There were involved 5 right wrists and 4 left wrists. All patients had a proximal pole of scaphoid non-union with necrosis and they failed the conservative or classical surgical treatment or result of diagnostic errors. The patients were submitted to preoperative MRI and TC, then the images were sent to ADLER^®^ company to create custom-made 3D implants. None of these patients had SNAC wrist or radio-carpal arthritis (Figs. 1 e 2). The approach was the dorsal-radial, with a sinusoidal incision of about 5 cm, medial to the course of the long extensor of the thumb, which was isolated and retracted, as well as the vascular structures and the sensory branches of the radial nerve. The radial extensors of the carpus have been mobilized and moved laterally to better highlight the radiocarpal dorsal ligament, which has been incised in inverted T-shape and dissected carefully by the carpus. All scaphoid bone was exposed and isolated. After removing the necrotic proximal pole and the remaining tissues of scapholunate ligament, the total distal pole was preserved. To reconstruct the scapholunate ligament, a palmaris longus tendon graft (PLTG) from the same wrist was used. Only in one case we had a problem to take palmaris longus tendon graft, so we chose to stabilize the implant with Garcia Elias’s tenodesis with FRC tendon graft. Then the implant was prepared to be placed. First, a tape of PLTG was fixed through the corresponding notch of the implant. The implant was fitted into its space, and the remaining tape of PLTG was stabilized with 1 or 2 anchors on the lunate (Fig.3). After confirming a satisfactory bx radiografically position (Fig.4), the stability of the implant was assessed by passively performing flexion and extension, laterality and rotation movements, before proceeding with the reconstruction of the radiocarpal dorsal ligament which was inserted on the radius with transosseous suture. Only in 3 cases we strengthened the ligament and the dorsal capsule with a flap of the extensor retinaculum. A suction drain was left in place for 24 h just to prevent hematoma formation. The day after surgery a thermoplastic splint was applied with the wrist in 10° extension. Immobilization was sustained for 4 weeks and then X-rays were performed to confirm the satisfactory position of the implant. Subsequent rehabilitation was then conducted for 3 months. Clinical and X-rays checks are performed at 1, 3, 6 and 12 months after the surgical intervention to follow the restoration of range of motion and to detect any early displacement of the implant or any changes in the carpal bones (Fig.5).

**Fig. 1:**
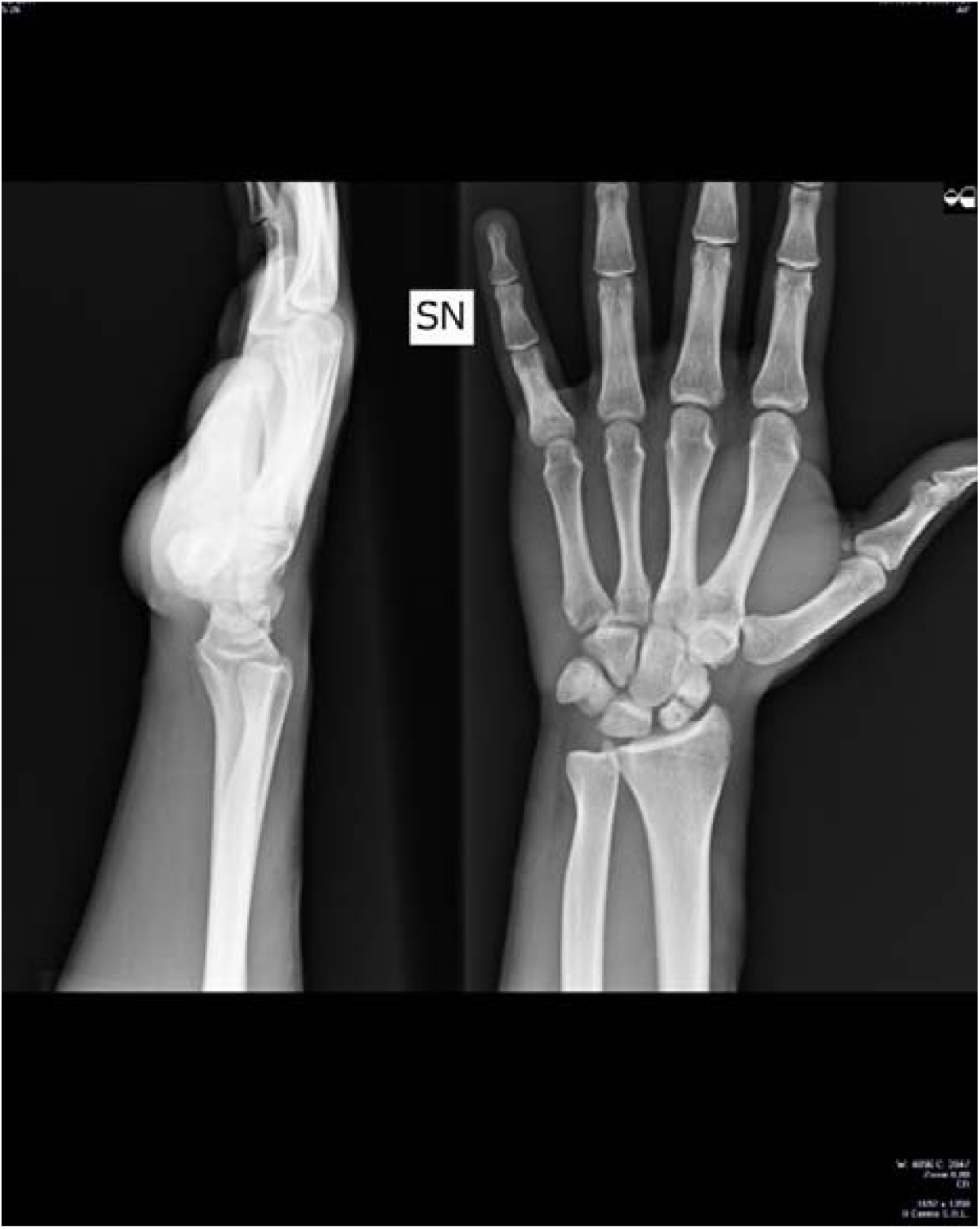
Typical pre-operative Xray image of a case of proximal pole of scaphoid nonunion. No evidence of SNAC wrist or radio-carpal arthritis

**Fig. 2:**
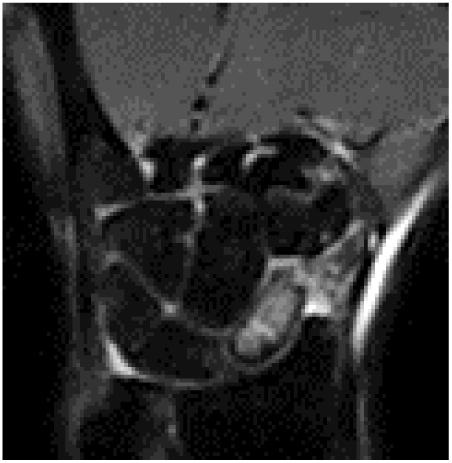
Typical pre-operative MRI image of a case of proximal pole of scaphoid nonunion with necrosis area.

**Fig. 3:**
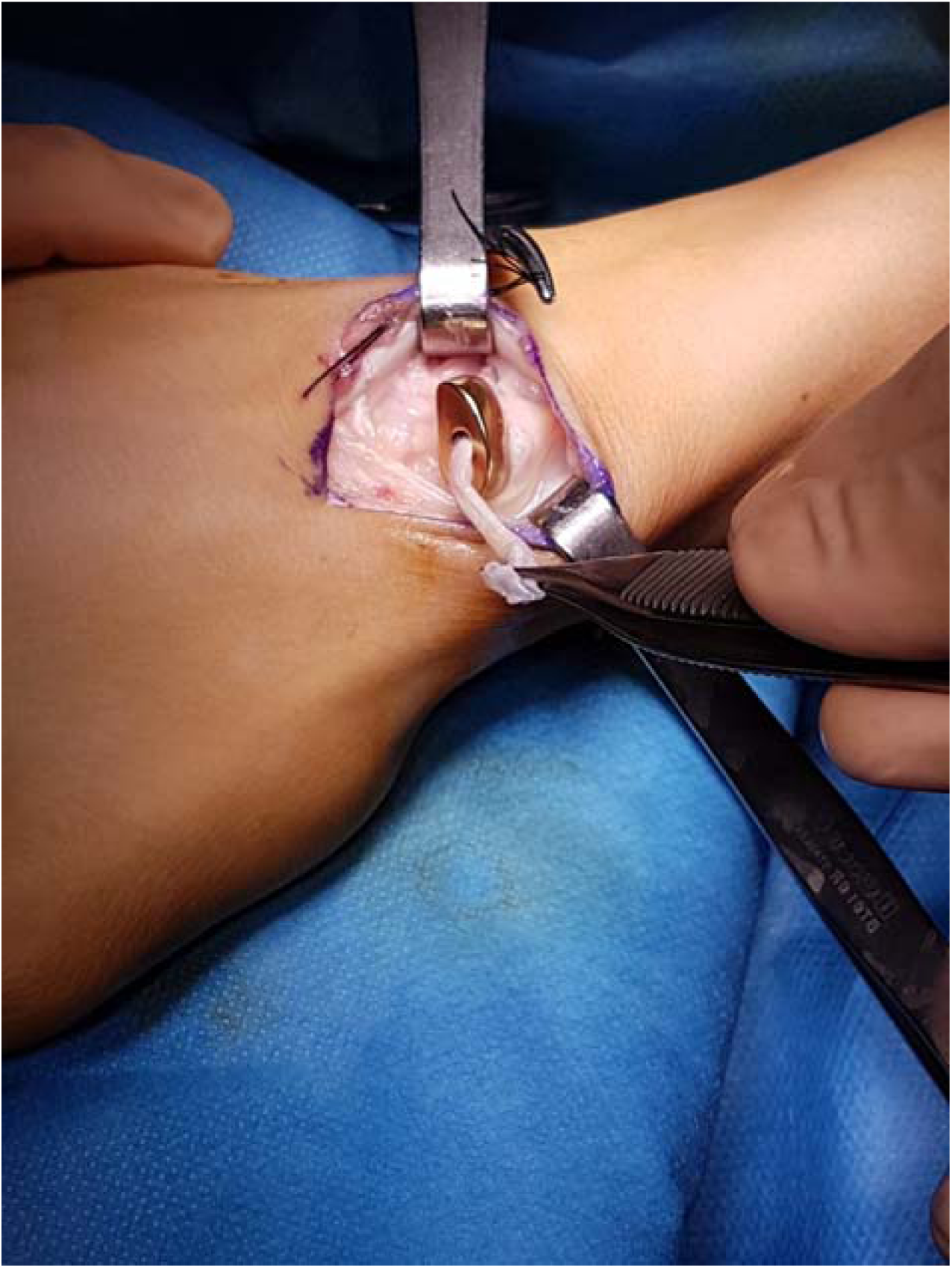
Intraoperative view. The titanium custom-made 3D-printed prosthesis is put in place; a PLTG passing through the prosthetic body will be fixed to the lunate.

**Fig. 4:**
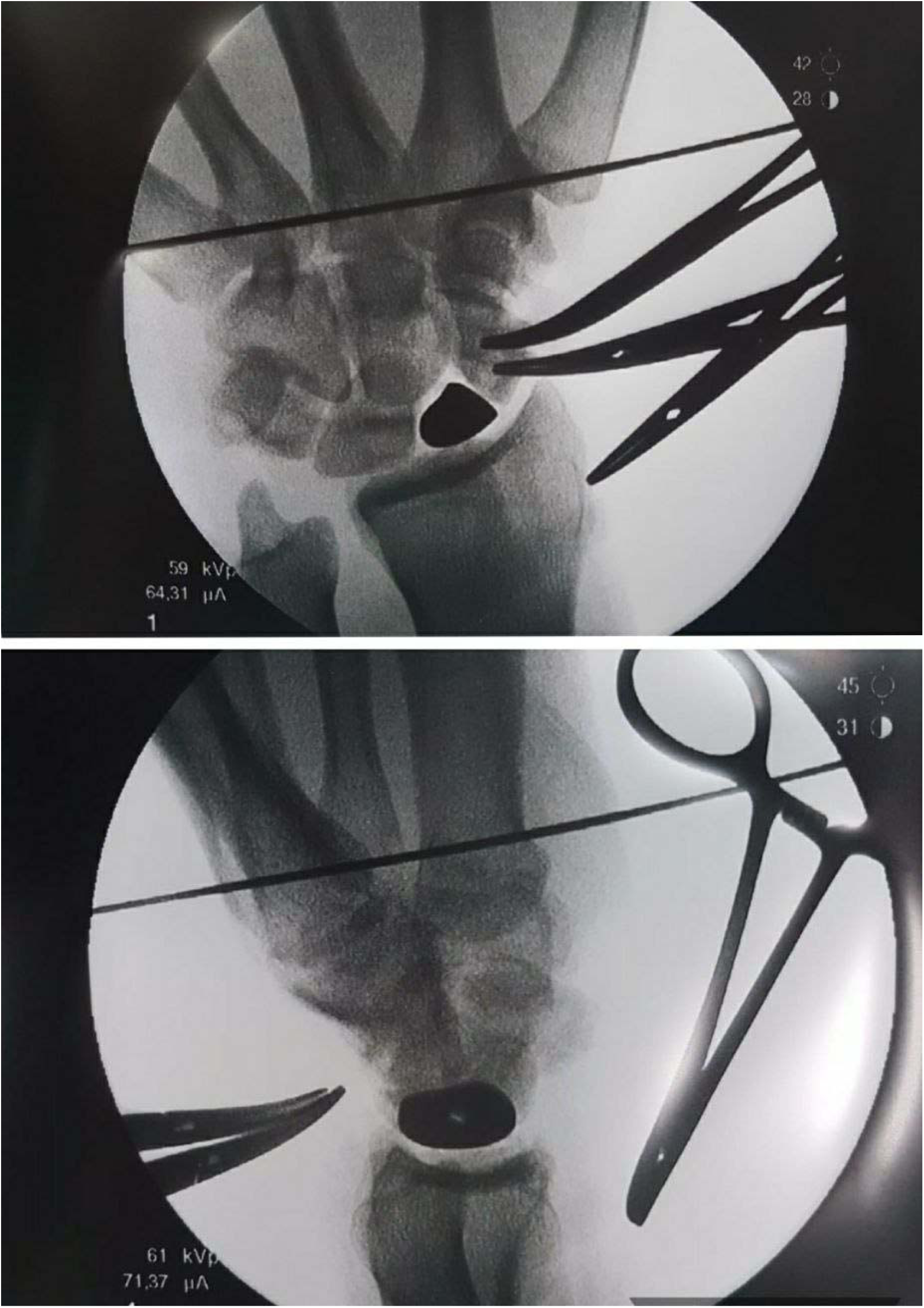
The perioperative radiography, the good prosthesis position was verified.

**Fig. 5:**
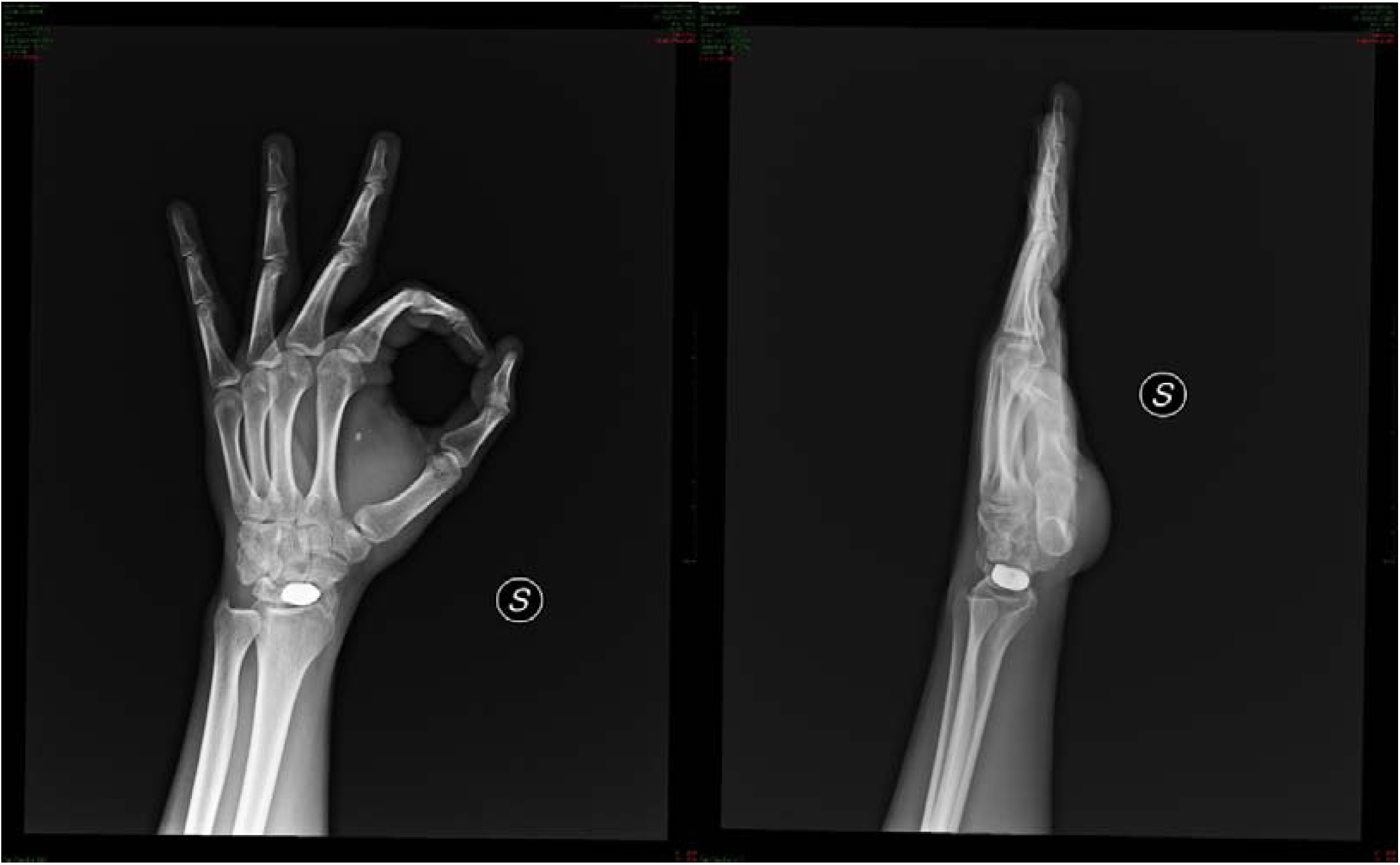
At 1-year follow up, radiography showed a correct radioscaphoid angle, no displacement or sign of carpal arthritis or other problems.

### Total scaphoid prosthesis

Between January 2019 and July 2020 in the Hand Surgery Department of San Paolo’s Hospital in Savona (Italy) 10 total scaphoid titanium custom-made 3D-printed prosthesis have performed in 10 patients (8 male and 2 female) with average age of 33.3 years (minimum 26-maximum 41 years). There were involved 4 right wrists and 6 left wrists. All patients had scaphoid non-union, as result of diagnostic error or failure of surgical treatments and irreparable bone destruction. However, they had good wrist stability, no SNAC wrist condition (as demonstrated by carpal height and radio-lunate angle measurements) and absence of STT arthritis. Patients were submitted to preoperative MRI and TC, then the images were sent to ADLER^®^ company to create a correct size of custom-made 3D implant. The approach was the same for the partial scaphoid prosthesis. When the capsule was opened, all necrotic scaphoid was removed, except a small, volar portion of the distal pole, of about 3×3 mm, must be left in place to preserve the radio-scapho-capitate ligament insertion. Then a hole was made in the trapezium under the control of the image amplifier. This hole was designed to host the distal tip of the prosthesis, a key point to stabilize the implant. To reconstruct the scapholunate ligament, Arthrex™ labral tape (Naples, FL, USA) was inserted into the lunate with an anchor. The implant was then prepared for the placement. First, the two cords of the labral tape were passed through the corresponding holes of the implant, and then the prosthetic distal tip was inserted into the trapezium hole^5^. The implant was fitted into its space, and the two labral tape cords were finally knotted in an apposite notch carved into the implant (Fig 6). The implant was therefore stabilized on both sides: distally by its tip inserted into the trapezium and proximally thanks to the labral wire fixed to the lunate. After confirming a radiographically satisfactory position, the implant stability was tested by moving the wrist in any direction. The closure of the capsule, the thermoplastic splint and postoperative checks were the same of partial prosthesis. Only in one case we had a distal dislocation of the stem at 1 month and we removed the implant and used 3-corner arthrodesis to stabilize the wrist.

**Fig. 6:**
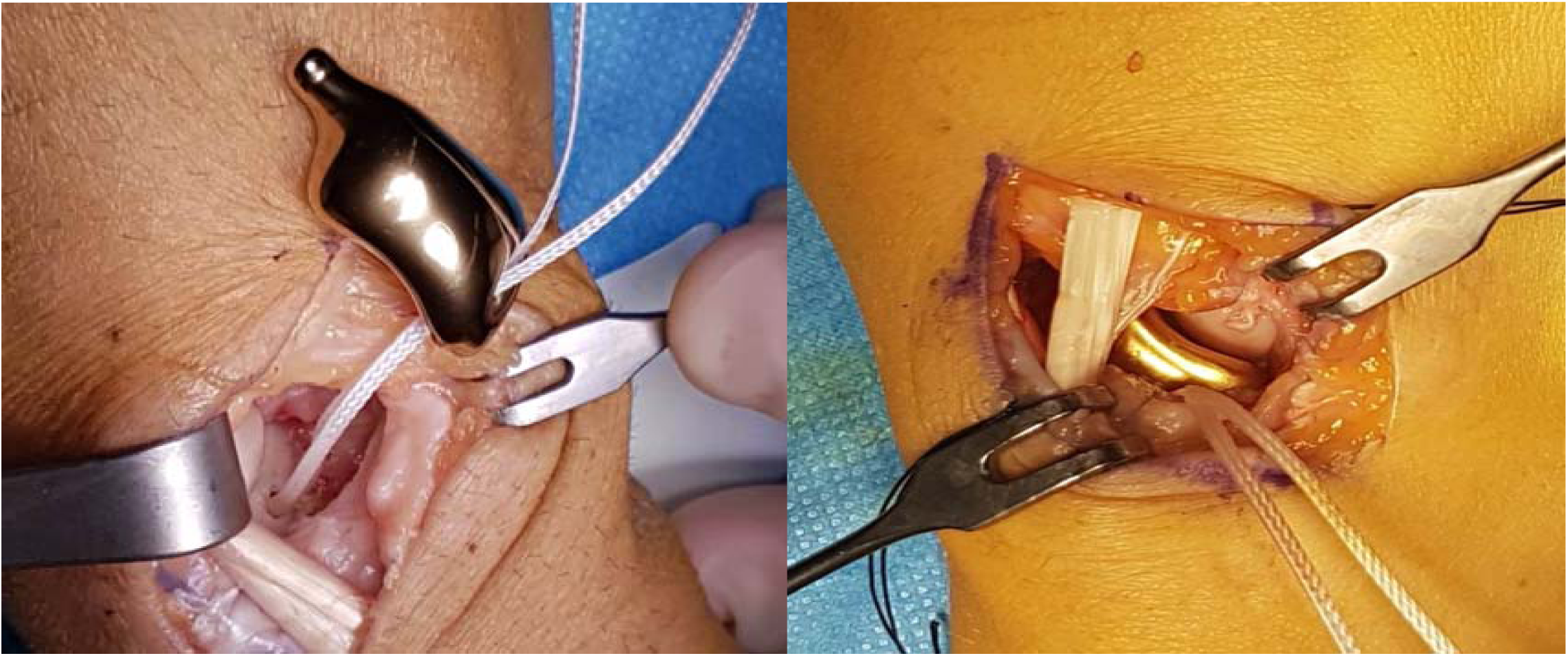
The titanium implant was positioned. ArthrexTM labral tape was fixed into the lunate and passed through the implant. The distal stem of the implant was set into the hole prepared into the trapezium.

**Fig. 7:**
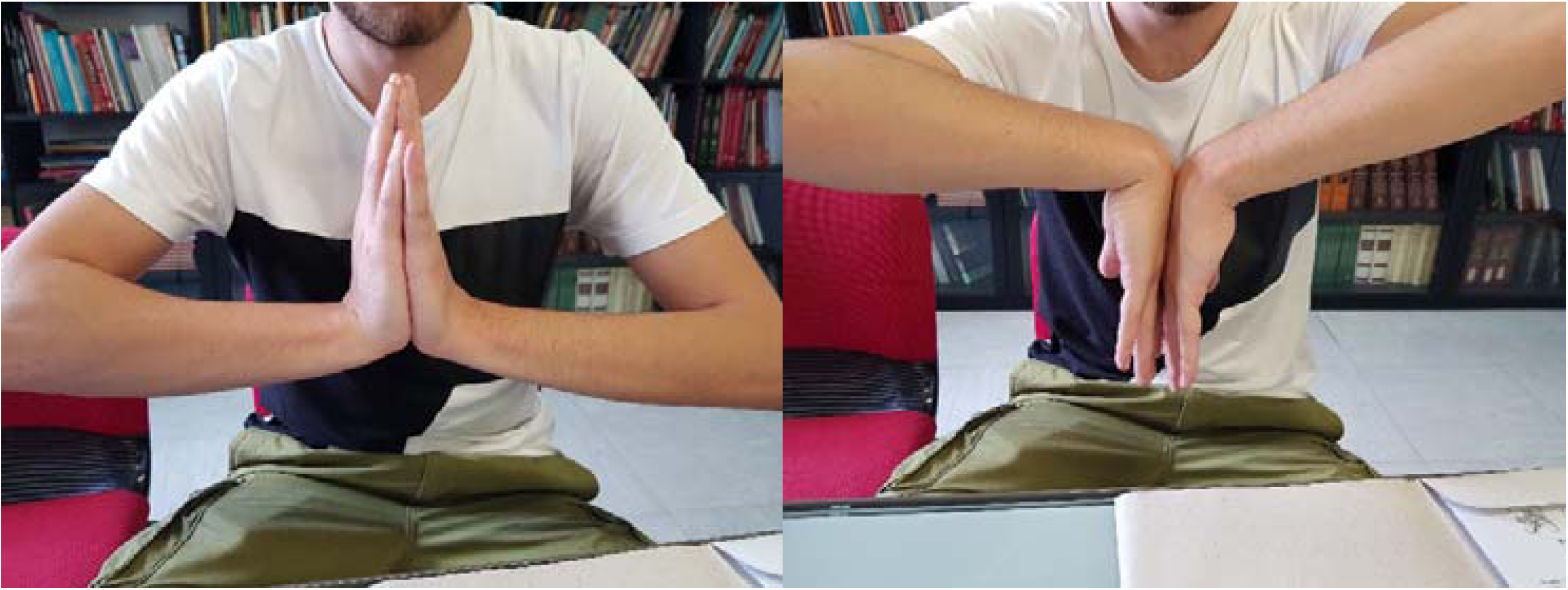
The range of motion after 1 year

## RESULTS

### Partial scaphoid prosthesis

We have been able to follow all the 9 patients with a mean follow-up of 18 months (minimum 12 months maximum 18 months). The evaluation criteria include carpal height ratio measured by Youm’s method, radioscaphoid angle, extension and flexion wrist, radial deviation and ulnar deviation of the wrist, grip strength and pinch strenght by Jamar test on operated wrist and contralateral, pain VAS scale at rest and under load, DASH score and patient-rated wrist evaluation (PRWE). We have evaluated DASH score and PRWE pre-operative and 1 year post operation and the other outcomes on pre-operative, 1 month, 3 months, 6 months and 12 months post-operative. The X-rays were useful to show the correct position of prosthetic stem into trapezium body, any evidence of reabsorption or cysts in the carpal bones and any sign of biocompatibility. The clinical findings (Table 1) demonstrated that the carpal height ratio decreased of 9,5% from pre-operative to 1 year. The radioscaphoid angle decreased of 29% at 1 year. Flexion and extension wrist increased about 34 % and 48% in 1 year from pre-operative. Radial deviation increased of 45% in 1 year, and ulnar deviation of 23 %. The grip strength improved from pre-operative to 1 year post-operative of 41,5%, the pinch strength of 38%. The VAS scale under load decreased of 77%. DASH score decreased about 59 % from pre-operative to 1 year, and PRWE about 48%. All these rates are an average. We hadn’t failure of the implant. We had only one failure in one case of the total prosthesis was dislocation at 1 month, and it was necessary to remove the implant and perform a 3-corner arthrodesis.

**Table 1:** Rx and Clinical outcomes of partial prosthesis

|  | PRE-OPERATIVE | 1 MONTH | 3 MONTHS | 6 MONTHS | 12 MONTHS |
| --- | --- | --- | --- | --- | --- |
| **CARPAL HEIGHT** | 0,84 (1,98-0,52) | 0,77 (1,96-0,45) | 0,77 (1,96-0,44) | 0,76 (1,95-0,44) | 0,76 (1,94-0,44) |
| **RADIOSCAPHOID ANGLE** | 45,75 ° (66-37) | 30,62 ° (43-20) | 32,5 ° (42-30) | 33,87 ° (43-30) | 32,37 ° (43-30) |
| **EXTENSION** | 33,57 ° (70-0) | 53,57 ° (65-30) | 57,14 ° (70-35) | 63,57 ° (80-50) | 64,28 ° (80-50) |
| **FLEXION** | 40 ° (70-20) | 41,42 ° (50-35) | 47,85 ° (60-40) | 59,28 ° (70-45) | 60,71 ° (70-45) |
| **RADIAL DEVIATION** | 12,85 ° (35-5) | 17 ° (22-5) | 21,42 ° (30-15) | 26,42 ° (60-15) | 26,45 ° (60-15) |
| **ULNAR DEVIATION** | 27,85 ° (45-10) | 36,14 ° (50-30) | 39,28 ° (45-35) | 37,14 ° (50-20) | 36,42 ° (50-20) |
| **GRIP (JAMAR TEST)** |  |  |  |  |  |
| **OPERATED W. CONTROLATERAL** | 17 kgm (22-10) | 12 kgm (15-10) | 15,6 kgm (24-10) | 24,7 kgm (31-20) | 29,1 kgm (38-24) |
|  | 39 kgm (50-26) | 37 kgm (46-25) | 37,1 kgm (48-26) | 36,4 kgm (48-26) | 30.8 kgm (50-28) |
| **PINCH** |  |  |  |  |  |
| **OPERATED W.** | 5,1 kgm (6,5-4) | 6,2 kgm (8,5-3,5) | 6,6 kgm (8,5-3,5) | 7,5 kgm (8,9-6) | 8 kgm (9-7) |
| **CONTROLATERAL** | 8,6kgm (10-6,5) | 8 kgm (9,5-6) | 8,6 kgm (9,5-6,5) | 8,5 kgm (9,5-6,5) | 8,6 kgm (9,5-6,5) |
| **PAIN VAS SCALE** |  |  |  |  |  |
| **AT REST** | 3,6 (10-1) | 1,9 (5-0) | 0,4 (1-0) | 0 | 0 |
| **UNDER LOAD** | 7,4 (10-5) | 4,4 (8-2) | 3,1 (5-2) | 2,5 (4-2) | 1,7 (3-1) |
| **DASH SCORE** | **PRE-OPERATIVE** |  | **1 YEAR** |  |  |
|  | 22,4 (51-12,9) |  | 9,2 (30,8-0,8) |  |  |
| **PRWE** | **PRE-OPERATIVE** |  | **1 YEAR** |  |  |
|  | 33,7 (88-28) |  | 17,5 (62-2) |  |  |

### Total scaphoid prosthesis

We were able to follow 10 patients with a median follow-up of 18 months (minimum 12 months maximum 18 months). The evaluation criteria were the same for the partial implant. The clinical results (Table 2) demonstrated that the carpal height ratio decreased of 13% from pre-operative to 1 year. The radioscaphoid angle decreased of 16% at 1 year. Flexion and extension of the wrist increased about 46 % and 40% in 1 year from pre-operative. Radial deviation increased of 65% in 1 year, and ulnar deviation of 16 %. The grip strength improved from pre-operative to 1 year post-operative of 47%, the pinch strength of 41%. The VAS scale under load decreased of 83%. DASH score decreased of 58 % from pre-operative to 1 year, and PRWE by 37%. All these rates are just an average. Of the total prosthesis we had just one failure in one case which was a dislocation at 1 month, and it was necessary to remove the implant and perform a 3-corner arthrodesis.

**Table 2:** Rx and Clinical outcomes of total prosthesis

|  | **PRE-**<br>**OPERATIVE** | **1 MONTH** | **3 MONTHS** | **6 MONTHS** | **12 MONTHS** |
| --- | --- | --- | --- | --- | --- |
| **CARPAL HEIGHT** | 0,61 (0,85-0,50) | 0,59 (0,80-0,45) | 0,58 (0,80-0,45) | 0,53 (0,80-0,45) | 0,53 (0,80-0,45) |
| **RADIOSCAPHOID**<br>**ANGLE** | 40,9 °(60,2-12) | 34,6° (58-44,5) | 35 ° (58-32) | 36,8 ° (58-30) | 34,2 ° (58-30) |
| **EXTENSION** | 31,4 ° (55-20) | 42 ° (65-25) | 47,8 ° (70-30) | 47,1 ° (70-35) | 52,1 ° (75-45) |
| **FLEXION** | 25,7 ° (45-10) | 36,7 ° (50-12) | 44,2 ° (60-20) | 44,2 ° (60-25) | 47,8 ° (60-35) |
| **RADIAL**<br>**DEVIATION** | 10 ° (25-0) | 13,5 ° (25-3) | 16,8 ° (25-8) | 20 ° (30-10) | 28,5 ° (45-20) |
| **ULNAR**<br>**DEVIATION** | 29,7 ° (60-10) | 29 ° (35-20) | 29,7 ° (35-25) | 32,1 °(45-30) | 35,5 ° (45-30) |
| **GRIP (JAMAR**<br>**TEST)** |  |  |  |  |  |
| **OPERATED W.** | 13,6 kg m (20-4) | 15 kg m (22-5) | 18,8 kg m (26-12) | 22,4 kg m (30-13) | 25,7 kg m(32-18) |
| **CONTROLATERAL** | 29 kg m (46-25) | 28,7 kg m (46-24) | 34,7 kg m (46-24) | 37,5 kg m (52-24) | 36,5 kg m(48-24) |
| **PINCH** |  |  |  |  |  |
| **OPERATED W.** | 5 kg m (10-1,8) | 4,7 kg m (12-2) | 10,5kgm(16-2,5) | 8 kg m (16-5) | 8,6kgm(16-10) |
| **CONTROLATERAL** | 11,2kgm(22-7) | 11,2kgm(22-6) | 10,5kgm(22-6) | 10,7kgm(22-6) | 10,7kgm(22-6) |
| **PAIN VAS SCALE** |  |  |  |  |  |
| **AT REST** | 5,5 (8-2) | 1,8 (3-1) | 1 (2-0) | 0,8 (2-0) | 0,1 (1-0) |
| **UNDER LOAD** | 8,2 (10-6) | 4,7 (8-2) | 3 (6-2) | 3 (5-0) | 1,4 (5-0) |
| **DASH SCORE** | **PRE-OPERATIVE** |  | **1 YEAR** |  |  |
|  | 28,2 (67,5-11,8) |  | 11,8 (28,3-1,7) |  |  |
| **PRWE** | **PRE-OPERATIVE** |  | **1 YEAR** |  |  |
|  | 35,6 (82-15) |  | 22,5 (51-7) |  |  |

## DISCUSSION

The treatment of scaphoid non-unions is still controversial because there are many options to restore the wrist cinematic which is based on the characteristics of each patient. The replacement of scaphoid with partial or total titanium custom made 3D printed prosthesis is a valid choice for young patients with an unsuitable scaphoid for a reconstruction with grafting techniques and without degenerative or arthritis signs in the wrist. We had an improvement in all analyzed outcomes, in partial and total prosthesis (Table 1 and 2). The best result was for VAS pain scale, with a great improvement from pre-operative to 1 year post-operative, at rest and under load. (Table 1 and 2). DASH score and PRWE was acceptable with a hight grade of satisfaction for the patients. The reconstruction of SLIL in both procedures allowed a greater stability and prevents a midcarpal bones collapse in the medium term. In case of failure, the same more aggressive procedures to be considered as alternatives are always possible, such as scaphoidectomy and 4- or 3-corner arthrodesis or proximal row carpectomy.^11^

## CONCLUSIONS

So far this study is the unique in Literature in this type of implants, even if preliminary. The use of these devices can mark new borders for the treatment of sequelae’s scaphoid fractures and an expression of the new technologies made available by orthopedic prosthesis companies and the maximum concept of “Custom-made Prosthesis” on anatomy of single patient. Several studies shown substantial inter-individual differences in the shape of the carpal bones and motion patterns of the scaphoid.^12-13-14^ Considering this we believe that any scaphoid prosthesis and any biological scaphoid reconstruction must replicate the original shape of the bone as precisely as possible to minimize non-physiological kinematics and wear. This concept requires a patient-specific implant: So far 3D printing is the only way to realize such a device. A strong pro for scaphoid implant reconstruction concept is that it doesn’t burn bridges, allowing in case of failure, the identical salvage procedures (PRC, 3 or 4 corner fusion) that could have been performed instead of. However, we believe that there are some critical issues for this study: the follow-up at only one year; the surgical approach must be dressed by expert hand surgery; the long time for manufacturing process of the prosthesis (about 3 weeks) and the elevate cost of the implants.

## Data Availability

This study was approved by our institutional review board.

